# MELODY trial: Study protocol for a 12-week randomised controlled trial of adjunctive melatonin, digital cognitive behaviour therapy for insomnia, or pill placebo to improve depressive symptoms in young adults with mood disorders

**DOI:** 10.1101/2025.09.08.25335055

**Authors:** Jacob J. Crouse, Mirim Shin, Samuel J. Hockey, Elie Jeon, Nathan Bradshaw, Alissa Nichles, Natalia Zmicerevska, Min Chong, Hannah You, Naomi R. Wray, Jan Scott, Ronald R. Grunstein, Sharon L. Naismith, Kathleen R. Merikangas, Sean W. Cain, Sarah E. Medland, Frank Iorfino, Thomas D.A. Uebergang, Sarah McKenna, Joanne S Carpenter, Emiliana Tonini, Nayonika Bhattacharya, Blake A Hamilton, Leanne M Wallace, Anjali K Henders, Elizabeth M. Scott, Yun Ju (Christine) Song, Brain and Mind Centre Lived Experience Working Group, Ian B. Hickie

## Abstract

**Objectives:** Sleep and circadian rhythm disturbances (SCRDs) are proposed to be pathophysiological mechanisms underlying some cases of depressive and bipolar (mood) disorders. An unresolved clinical question is whether sleep and circadian based therapies are effective antidepressants for young adults (18-30-years) with a mood disorder.

**Method & analysis:** MELODY (Melatonin for Depression in Youth) is an investigator-initiated, single-centre, randomised, placebo-controlled, phase 3 clinical trial. The trial is testing whether 12 weeks of adjunctive melatonin or digital cognitive behavioural therapy for insomnia (dCBT-I) are more effective than pill placebo at reducing depressive symptoms in 660 young people aged 18-30-years with a Structured Clinical Interview for DSM-5 (SCID-5) diagnosis of Major Depressive Disorder or Bipolar Disorder type II, moderate-to-severe depressive symptoms, and significant sleep or sleep-wake complaints. The week 12 primary outcome is depressive symptoms (Quick Inventory of Depressive Symptomatology, Adolescent version). Secondary outcomes include partial remission of the Major Depressive Episode (SCID-5) and change in other mental health symptoms, sleep, functioning, or quality of life. A subset of participants will undergo in-home assessment of sleep physiology (Sleep Profiler^TM^) and in-lab assessment of circadian rhythms (dim-light melatonin onset) to examine biological changes. A 6-month follow-up will explore durability of treatment effects. Mediation analyses will test whether sleep or circadian rhythm changes play a causal role in antidepressant effects.

**Ethics & dissemination:** MELODY has been reviewed and approved by the Human Research Ethics Committee (HREC) of the Sydney Local Health District (HREC Approval Number: X23-0450, Protocol version: 1.4, 7/7/25). The findings of the MELODY trial will be disseminated into the scientific and clinical communities via refereed publications, talks, and other professional and media outlets. The Brain and Mind Centre’s Lived Experience Working Group (LEWG) will contribute to dissemination of MELODY’s findings via youth-friendly methods (e.g., social media videos, explainers).

**Trial registration number:** ACTRN12624000017527

**Strengths and limitations of the study:**

- MELODY is the first trial to be well-powered (>80%) to detect the summary effect size reported in the only meta-analysis on the effect of melatonin on depressive symptoms in people with mood disorders (standardised mean difference=0.37); MELODY will therefore be the first definitive trial on the topic.
- As melatonin and digital cognitive behavioural therapy for insomnia (dCBT-I) are scalable treatments, the success of either arm compared to placebo is likely to lead to real-world clinical impacts.
- By recruiting a sample with a spectrum of sleep and sleep-wake complaints, post-hoc analyses may be able to identify profiles or subgroups that can guide stratified trials or guidelines for melatonin and dCBT-I in young people with mood disorders.
- While we expect participants in the dCBT-I arm will learn sustainable skills to improve their sleep, sleep-wake cycles, and circadian rhythms, the likelihood of durable gains from melatonin is unclear; this will be explored at 6-month follow-up.
- This study adopts a primarily biomedical and clinical approach to the understanding and treatment of mood disorders and therefore does not encompass all perspectives on mood disorders (e.g., social, cultural).

## INTRODUCTION

Improving the therapeutic landscape for young people with depressive and bipolar (mood) disorders is a global priority^1–7^. The peak phase of onset of mood disorders is during adolescence and early adulthood^5,8,9^. This is a time in which these conditions exert their most profound and enduring impacts on engagement with employment, education, and relationships^10–12^. Moreover, death by suicide^13,14^ and the premature morbidity caused by comorbid physical illnesses^15^ results in an average life expectancy 10-15 years lower than the general population^16^.

Despite the magnitude of illness and disability caused by mood disorders, the average treatment outcomes for these conditions are sobering^17,18^. For example, in the case of depressive disorders, a meta-analysis of over 1M people estimated that globally only 35% of individuals receive treatment^19^. Among those who *are* treated, only 40% receive minimally adequate treatment^19^. Trials testing safe, affordable, and potentially scalable treatments are needed, particularly for young people^2^, who have been the focus of fewer trials compared to middle-aged and older populations.

Sleep and circadian rhythm disturbances (SCRDs) have been proposed as targets for interventions for people with mood disorders^20–24^. There is compelling evidence from epidemiologic and interventional studies that sleep disturbances play a critical role in the onset and course of mental disorders^25–27^. There is suggestive (but nonconclusive) evidence of a critical role for circadian disturbances^23^, at least for a subgroup of people with mood disorders^21,28–31^. For example, in some studies, circadian disturbances are associated with the onset of mood disorders^32,33^, stage of illness (cross-sectionally)^34^, and progression to more severe stage of illness longitudinally^35^. If SCRDs are causative pathophysiological mechanisms of some types of mood disorders, then their correction may have therapeutic value. Two potential scalable interventions for SCRDs in young people with mood disorders are melatonin and digital cognitive behaviour therapy for insomnia.

Melatonin is a hormone produced by the brain’s pineal gland. It plays a key role in the timing of the 24-hr circadian system as the *chemical signal of darkness*^36^. Exogenous melatonin has long been demonstrated to have an effect on the phase of the circadian rhythm of melatonin^37,38^. However clinical trials investigating the efficacy of adjunctive melatonin for depressive symptoms in the context of mood disorders have yielded mixed (but nonconclusive) results. We are aware of nine trials examining the effect of melatonin on depressive symptoms in people with mood disorders (including unipolar, bipolar, and seasonal mood disorders)^39–47^. The first trial, published ∼50 years ago, reported that melatonin worsened depressive symptoms in six people with moderate-to-severe depression^39^—this was the only trial to report a *worsening* in the depressive state. Since then, six trials have reported that melatonin (either alone or as an adjunct to buspirone^40^) had no effect on depressive symptoms^41–44,47,48^. Three trials have reported that melatonin improved depressive symptoms^45,46,48^. A 2017 random-effects meta-analysis^49^ of three trials (pooled N=113) reported a non-significant effect of melatonin on depressive symptoms in people with mood disorders (SMD=0.39; 95% CI [−0.05 to 0.78]; P=0.09). While non-significant, this point estimate of 0.39 SD is substantial and could imply low statistical power given that mood disorders are heterogeneous. A systematic review by the *International Society for Bipolar Disorders* (ISBD) *Chronobiology and Chronotherapy Task Force* concluded in 2019 that there was insufficient or conflicting data regarding melatonin or melatonergic agents for the acute or maintenance treatment of depression in bipolar disorders^20^. A Delphi expert consensus study by the same Task Force (led by the first-author of this paper) concluded this is still the state of the evidence 5 years later^50^.

Each of these trials on exogenous melatonin contain methodological limitations that hamper the drawing of firm conclusions. The principal flaw is their small sample sizes, which range from 6-142 participants. Only one trial^40^ was adequately-powered (80%) to detect a one-tailed Cohen’s d effect size of ≤0.5 (the upper end of which is likely to be an inflated average effect size^49^). A second flaw is the common lack of selecting patients with significant SCRDs for whom it can be expected that melatonin would work for more effectively. To our knowledge, only three of the trials used such selection: one negative trial selected cases with major depressive disorder with concurrent early-morning wakening^43^, and two positive trials selected cases with a seasonal pattern of winter depression^45,46^. Altogether, we interpret this evidence base as non-conclusive. We conclude that a well-powered, randomised controlled trial is needed to *definitively* establish the efficacy of melatonin for the depressed phase of mood disorders in young people, and to examine whether it performs better in cases with significant sleep, sleep-wake, and/or circadian disturbances (or a specific profile).

Comparison of circadian versus sleep targeted treatments may help identify treatment relevant subgroups for stratified treatment models. The sleep therapy with the strongest evidence base is cognitive behaviour therapy for insomnia (CBT-I), which is a first-line approach to treating insomnia. Its effectiveness is supported by dozens of clinical trials. Digital CBT-I (dCBT-I) programs have been developed to enhance its accessibility, and dCBT-I has also been shown to be highly effective for the treatment of insomnia. Given the identification of insomnia as a probable causal mechanism of some forms of depressive symptoms and disorders^25^, exploration of the effect of dCBT-I on depressive symptoms (acting via improvement in sleep) has been of great interest^51–53^. A recent meta-analysis of 14 studies across several health conditions reported a small-to-medium beneficial effect of fully automated dCBT-I (i.e., no therapist support) on depressive symptoms (SMD = −0.43; 95% CI [−0.61 to −0.26]; P<0.001)^54^. Another meta-analysis of seven trials of dCBT-I in people with comorbid insomnia and depression reported a medium-to-large effect of dCBT-I on depressive symptoms (SMD = −0.60; 95% CI [−0.79 to −0.40]; P<0.001)^55^. The mean age of many samples in these meta-analyses^55^ are predominantly middle-aged (age 40-50) and only a handful of trials have been conducted in younger mood disorder populations^56^.

### Objectives of the study

There is a need for definitive trials of scalable sleep and circadian therapies in young mood disorder samples^21,23,57^ and head-to-head comparison of therapies with different mechanisms of action on SCRDs. The outcomes of such comparisons may be used to identify which therapies work best for whom^23,57^, particularly in the context of future stratified treatment models^21^ for people with mood disorders.

Accordingly, the main objective (primary planned analysis) of the MELODY trial is to investigate whether sleep and circadian targeted therapies (dCBT-I and melatonin) are more effective than placebo medicine at reducing depressive symptoms in young people with mood disorders. Other objectives include testing whether melatonin is more effective than dCBT-I at reducing depressive symptoms; whether improvements in sleep and circadian factors mediate reductions in depressive symptoms; and whether there are particular subgroups or profiles that respond preferentially to melatonin or dCBT-I. The outcomes of the MELODY trial will have significant implications for early intervention for young people with mood disorders, and for the causal status of SCRDs in these conditions (under the *interventionist* causal model^58^).

## METHODS & ANALYSIS

This protocol paper is written according to the 2013 SPIRIT guidelines^59^.

### Patient and public involvement

Members of the Brain and Mind Centre Lived Experience Working Group (LEWG)^21,60^ were consulted during the design and planning phases of the trial, and a Lived Experience Researcher is a Chief Investigator on the trial’s funding (227089/Z/23/Z). The LEWG consists of culturally and linguistically diverse adolescents and young adults aged 16-30 years. To date the LEWG have been engaged on the MELODY trial regarding eligibility criteria (e.g., decision to not exclude individuals who have taken melatonin previously); logo design; recruitment strategies; reimbursement rates; modes of communication with the study team; cultural considerations (e.g., timing study activities around periods such as Ramadan); and consent procedures (e.g., providing photographs of the overnight chronobiology suite). Throughout the lifetime of the trial, LEWG members will be engaged regularly to provide feedback on procedures, progress, and dissemination. Members are reimbursed in line with the Australian *Paid Participation Policy*, and consultation with the LEWG was approved by the University of Sydney’s Human Research Ethics Committee.

### Trial design, setting, and oversight

The MELODY trial is an investigator-initiated, three-arm, randomised, placebo-controlled, phase 3, double-blinded, superiority trial. The MELODY trial is conducted at the Brain and Mind Centre, University of Sydney, NSW, Australia.

A monitor that is independent from both the investigators and the trial sponsor has conducted a trial initiation visit and will conduct quarterly data monitoring audits, for-cause visits, and a trial close-out visit. The trial will be audited according to the Good Clinical Practice guidelines by the International Council for Harmonization.

An independent Data Monitoring and Ethics Committee (DMEC) comprised of three psychiatrists and one statistician, and an independently-chaired and majority-independent Trial Steering Committee (TSC), are appointed to oversee the trial.

### Trial population and inclusion and exclusion criteria

The trial is enrolling young adults in Sydney, Australia, who have a unipolar major depressive disorder or bipolar disorder type II (BD-II), and who report significant difficulties with their sleep and/or sleep-wake cycle. Potential participants can be referred by their clinician or they can self-refer to the study.

#### Inclusion criteria

- Aged 18-30 (inclusive at the time of providing consent); and
- Current diagnosis of Major Depressive Disorder (MDD) or Bipolar Disorder type II (BD-II), confirmed via the Structured Clinical Interview for DSM-5 (SCID-5)^61^ at the time of enrolment; and
- A score of ≥11 on the Quick Inventory of Depressive Symptomatology, Adolescent version, Clinician Rating (QIDS-A-CR)^62^, indicating a moderate-to-severe Major Depressive Episodes (MDE), assessed within 14 days prior to study entry; and
- Evidence of significant sleep or sleep-wake complaints, indicated by any of the following:

`o` Moderate-to-severe sleep disturbance, indicated by a T-score ≥ 60 on the Patient-Reported Outcome Measurement Information System (PROMIS^TM^) Sleep Disturbance 8-item short form^63^; or
`o` Sleep-related impairment, indicated by a T-score ≥ 60 on the Patient-Reported Outcome Measurement Information System (PROMIS^TM^) Sleep-Related Impairment 8-item short form^63^; or
`o` Irregular sleep-wake timing, according to a response of “Never” or “Rarely” to the following modified item from the RU-SATED^64^ scale, “Generally speaking, I go to bed and get out of bed at about the same time every day (within 2 hours), disregarding changes due to social occasions”; or
`o` Delayed sleep phase, indicated by self-reported habitual sleep onset time ≥ 2 hours later than the desired or required bedtime; and
- Ability to provide written informed consent, including both adequate intellectual capacity and fluency in English

#### Exclusion criteria

- Use of any form of melatonin or melatonin agonist within six weeks prior to study entry; or
- Initiation of new medication to treat a mental health condition within four weeks prior to study entry; or
- Current use of a wakefulness-promoting medication, such as Modafinil; or
- Current use of Warfarin
- Acute suicidal behaviour, indicated by a score of 6 or more on the Comprehensive Assessment of At-Risk Mental States (CAARMS)^65^ item 7.3; or
- Self-reported diagnosis or treatment for a psychotic syndrome; or
- Self-reported diagnosis or treatment for an alcohol or substance use disorder; or
- Self-reported diagnosis or treatment for significantly impaired kidney or liver function; or
- Self-reported diagnosis or treatment for major sleep, respiratory or neurological

### disorders, or other medical condition causing significant sleep-wake dysfunction; or

- Self-reported allergy to melatonin or Microcrystalline Cellulose; or
- Self-reported pregnancy or breastfeeding; or
- Participation in another trial of a sleep or circadian therapy within four weeks prior to study entry; or
- Regular night shift work within two months prior to study entry; or
- Recent transmeridian travel, defined as crossing two or more time zones within four weeks prior to study entry; or

### Interventions

#### Melatonin and placebo

Participants allocated to a medication arm will receive a code-labelled bottle containing capsules of either melatonin (2mg) or placebo, to be taken each night, two hours before their desired bedtime. At week 4 (and subject to an assessment by the Study Doctor) the dose will be escalated to two capsules of melatonin (4mg) or placebo. Participants in the medication arms will be provided with a brief instructional video wherein the Principal Investigator (I.B.H.) discusses the rationale and procedure for taking melatonin, the importance of concurrently making relevant behavioural modifications (e.g., decreasing exposure to artificial light at night, increasing exposure to natural daylight), and instructions to try to align their sleep period within the dark period of the day (between ∼9pm-7am).

#### Digital CBT for insomnia (dCBT-I)

Participants allocated to the digital arm will be given access to ‘Sleep Healthy Using the Internet’ (SHUTi), an interactive, internet-based CBT-I intervention^66^. The program includes six sequential, interactive “core” modules, each of which take 45-60 minutes to complete; each new “core” module becomes available one week after completion of the previous core. Participants will be instructed by study staff to complete one module per week and to implement what they have learned during the remaining 12-week intervention period. They will be encouraged to return to previously completed modules as much as they would like to consolidate their learnings. The six “core” modules cover the definition of insomnia and its impact and risk factors; the rationale for dCBT-I in treating insomnia; sleep restriction and stimulus control strategies to strengthen the homeostatic sleep drive and increase sleep efficiency; education about good sleep and sleep hygiene practices (e.g., increasing exercise, avoiding meals and stimulants around bedtime); cognitive restructuring to challenge unhelpful beliefs and thoughts about sleep; and strategies to promote adherence to what has been learned through the program and strategies to prevent insomnia relapse. Participants in the trial will have access to a personalised home page showing their progress through SHUTi, and feedback and personalised tailoring of SHUTi is based on information provided by participants in an online daily sleep diary.

#### Treatment as Usual (TAU)

All participants will continue to receive any TAU from their treating clinician, which is not considered part of the trial activity.

#### Adverse events (AEs)

At baseline, a detailed self-report assessment of AEs common to melatonin and antidepressant medications (for the latter, using the *Antidepressant Side-Effect Checklist* ^67^) will be collected before the intervention. Thereafter, regular assessment of AEs will be conducted. Information about ancillary and post-trial care and withdrawal/discontinuation is in the Appendix.

#### Adherence

Participants in the medication arms will be asked to complete a medication diary, and the study team will remotely monitor adherence to SHUTi.

#### Reimbursement

Information about reimbursement is in the Appendix.

### Randomisation

Participants are randomly assigned in a 1:1:1 ratio to receive melatonin, placebo medicine, or dCBT-I in variable block sizes, with stratification according to age (dichotomised as 18-24-years vs 25-30-years), sex assigned at birth (male vs female), and baseline severity of depressive symptoms (*Quick Inventory of Depressive Symptomatology Clinician Rated Adolescent Version* [QIDS-A-CR] score dichotomised as 11-15 [moderate] vs 16+ [severe to very severe]). The randomisation sequence was generated electronically by a statistician not affiliated with the trial and transferred to the research team’s trial database (REDCap^68^). The drug manufacturer created labels on pill bottles indicating the medication arm (melatonin vs placebo) which are torn off by an unblinded member of the trial team during dispensation. Trial participants and assessors are blinded to medication allocation (melatonin vs placebo) but not to allocation of medication vs dCBT-I. Information about unblinding is in the Appendix.

### Sample size calculation

Meta-analyses report an effect size of 0.37 for melatonin (vs placebo) on depressive symptoms^49^ and 0.35 for dCBT-I (vs various comparators) on depressive symptoms^69^. A minimum effect size of interest of 0.30 is clinically meaningful and a feasible target. Assuming an attrition rate of 20% based on RCTs in a similar population^70^ (note: a lower attrition rate of 16% has been reported for academia-led pharmacological mental health trials^71^), 660 participants (220 per arm) are needed to provide 80% power to detect differences in change from baseline (d=0.3) at α=0.05 (two tailed test).

### Study course and procedures

All trial activities will be coordinated from the University of Sydney’s Brain and Mind Centre (BMC). A summary of the course and procedures of the trial is in Figure 1 and Table 1, and the assessments are summarised in Table 2. Advertisements for the trial will be distributed via social media, engagement with clinical contacts (e.g., *headspace*, *Mind Plasticity*), and flyers given to clinics and distributed around local universities, with a QR code leading to the study website (hosted on REDCap^68^). The study website contains a plain language summary about the study procedures and a copy of the consent form (PIS/CF). Upon completion of a brief online pre-screening form, potential participants will be contacted to schedule an in-person enrolment visit (Table 1).

**Figure 1.**
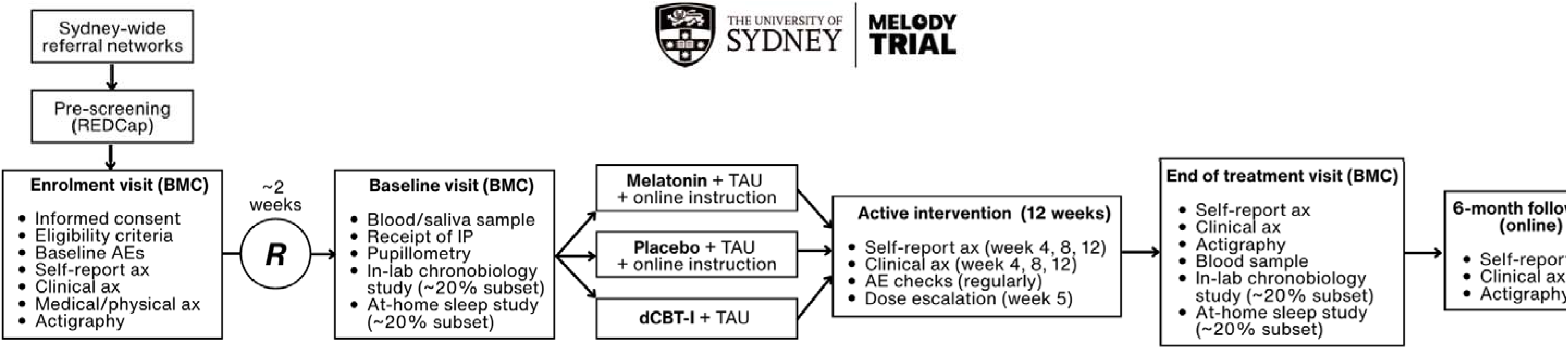
MELODY trial schematic.

**Table 1.**
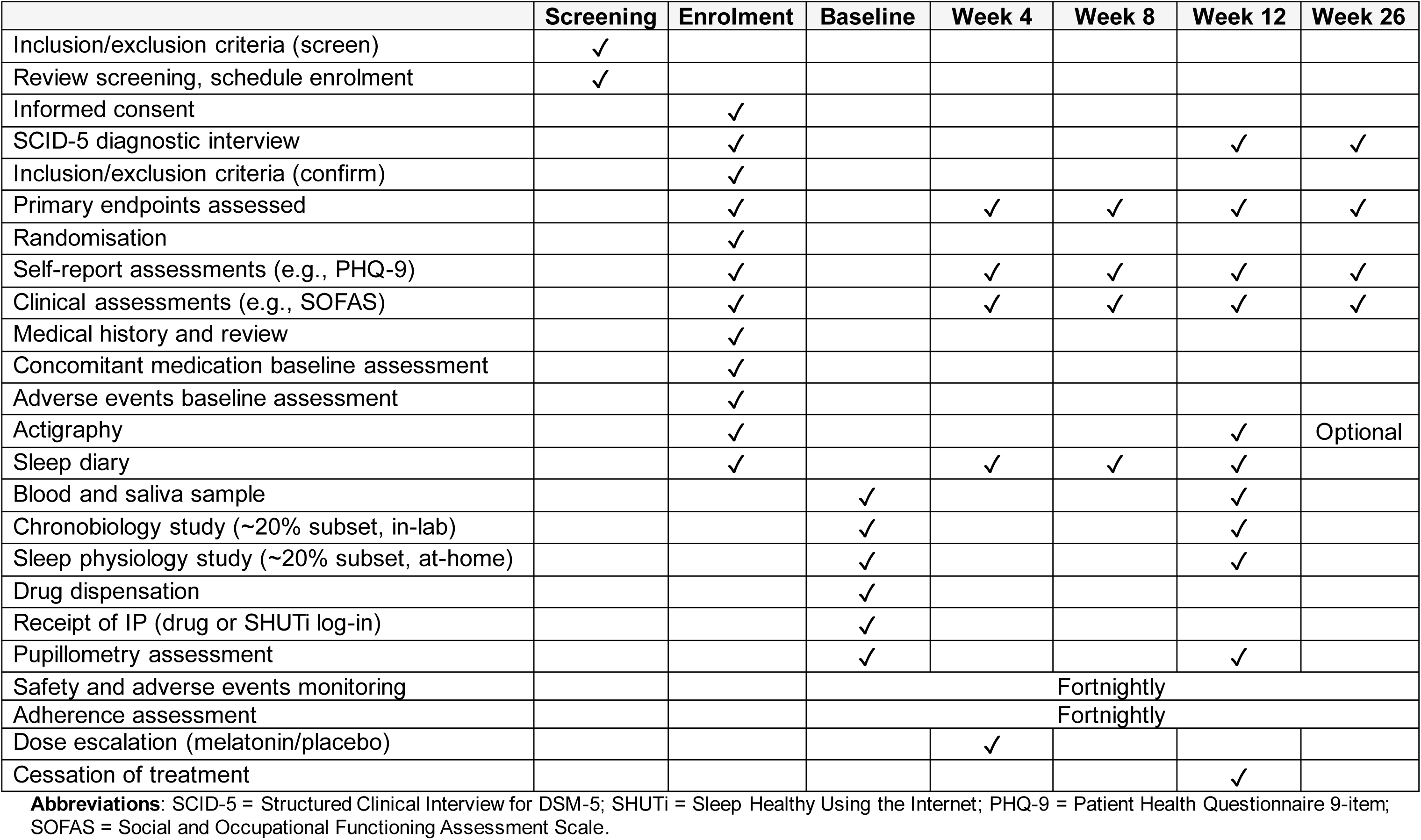
Overview of study course and procedures.

|  | Screening | Enrolment | Baseline | Week 4 | Week 8 | Week 12 | Week 26 |
| --- | --- | --- | --- | --- | --- | --- | --- |
| Inclusion/exclusion criteria (screen) | ? |  |  |  |  |  |  |
| Review screening, schedule enrolment | ? |  |  |  |  |  |  |
| Informed consent |  | ? |  |  |  |  |  |
| SCID-5 diagnostic interview |  | ? |  |  |  | ? | ? |
| Inclusion/exclusion criteria (confirm) |  | ? |  |  |  |  |  |
| Primary endpoints assessed |  | ? |  | ? | ? | ? | ? |
| Randomisation |  | ? |  |  |  |  |  |
| Self-report assessments (e.g., PHQ-9) |  | ? |  | ? | ? | ? | ? |
| Clinical assessments (e.g., SOFAS) |  | ? |  | ? | ? | ? | ? |
| Medical history and review |  | ? |  |  |  |  |  |
| Concomitant medication baseline assessment |  | ? |  |  |  |  |  |
| Adverse events baseline assessment |  | ? |  |  |  |  |  |
| Actigraphy |  | ? |  |  |  | ? | Optional |
| Sleep diary |  | ? |  | ? | ? | ? |  |
| Blood and saliva sample |  |  | ? |  |  | ? |  |
| Chronobiology study (~20% subset, in-lab) |  |  | ? |  |  | ? |  |
| Sleep physiology study (~20% subset, at-home) |  |  | ? |  |  | ? |  |
| Drug dispensation |  |  | ? |  |  |  |  |
| Receipt of IP (drug or SHUTi log-in) |  |  | ? |  |  |  |  |
| Pupillometry assessment |  |  | ? |  |  | ? |  |
| Safety and adverse events monitoring |  |  | Fortnightly |  |  |  |  |
| Adherence assessment |  |  | Fortnightly |  |  |  |  |
| Dose escalation (melatonin/placebo) |  |  |  | ? |  |  |  |
| Cessation of treatment |  |  |  |  |  | ? |  |

**Abbreviations:** SCID-5 = Structured Clinical Interview for DSM-5; SHUTi = Sleep Healthy Using the Internet; PHQ-9 = Patient Health Questionnaire 9-item; SOFAS = Social and Occupational Functioning Assessment Scale.

**Table 2.** Overview of research assessments.

| Domain | Assessment | Administration | Time points (weeks) |  |  |  |  |
| --- | --- | --- | --- | --- | --- | --- | --- |
|  |  |  | 0 | 4 | 8 | 12 | 26 |
| Demographics | Standard items | Self-report | ? |  |  |  |  |
| Medical history and review | Standard items | Researcher | ? |  |  |  |  |
| Clinical diagnosis | Structured Clinical Interview for DSM-5 | Researcher | ? |  |  | ? | ? |
| Clinical staging | Transdiagnostic staging model<sup>85</sup> | Researcher | ? |  |  | ? | ? |
| Illness trajectory | BMC illness trajectory model<sup>1</sup> | Researcher | ? |  |  |  |  |
| Acute suicidal behaviour (exclusion criteria) | CAARMS (7.3) | Researcher | ? | ? | ? |  |  |
| Depressive symptoms | QIDS-A-CR | Researcher | ? | ? | ? | ? | ? |
| Depressive symptoms | QIDS-A-SR | Self-report | ? | ? | ? | ? | ? |
| Depressive symptoms | PHQ-9 | Self-report | ? | ? | ? | ? | ? |
| Depressive and anxiety symptoms | RCADS | Self-report | ? | ? | ? | ? | ? |
| Anxiety symptoms | GAD-7 | Self-report | ? | ? | ? | ? | ? |
| Anxiety symptoms | OASIS | Self-report | ? |  |  | ? | ? |
| Insomnia symptoms | ISI | Self-report | ? | ? | ? | ? | ? |
| General sleep disturbance | PROMIS Sleep disturbance (short-form) | Self-report | ? | ? | ? | ? | ? |
| General sleep-related impairment | PROMIS Sleep-related impairment (8a) | Self-report | ? | ? | ? | ? | ? |
| Suicidal thoughts and behaviours | SIDAS | Self-report | ? | ? | ? | ? | ? |
| Morningness-eveningness (chronotype) | MEQ | Self-report | ? |  |  | ? | ? |
| Social and occupational functioning | SOFAS | Researcher | ? |  |  | ? | ? |
| Hypo/manic symptoms | ASRM | Self-report | ? |  |  | ? | ? |
| Disability | WHODAS-2.0 | Self-report | ? |  |  | ? | ? |
| Quality of life | ReQoL | Self-report | ? |  |  | ? | ? |
| Alcohol and substance use | WHO-ASSIST | Self-report | ? |  |  | ? | ? |
| Common side-effect symptoms | ASEC | Self-report | ? |  |  | ? |  |
| Physical health | Standard clinical bloods (optional) | Researcher | ? |  |  | ? |  |
| Physical health | Vital sign assessment | Researcher | ? |  |  | ? |  |
| Genetic risk (DNA) | Saliva sample | Objective | ? |  |  |  |  |
| Sleep-wake cycles | Actigraphy | Objective | ? |  |  | ? | ? |
| Circadian rhythms | Salivary melatonin sampling (DLMO) | Objective | ? |  |  | ? |  |
| Sleep physiology | Sleep Profiler™ EEG Sleep Monitor | Objective | ? |  |  | ? |  |
| Light sensitivity | Pupillary light reflex (PLR-3000) | Objective | ? |  |  | ? |  |

**Abbreviations:** CAARMS=Comprehensive Assessment of At-Risk Mental States; QIDS-A-CR=Quick Inventory of Depressive Symptomatology (Adolescent), clinician-rated; QIDS-A-SR=QIDS, self-report; PHQ-9=Patient Health Questionnaire 9-item; RCADS=Revised Children's Anxiety and Depression Scale; GAD-7=Generalized Anxiety Disorder 7-item; ISI=Insomnia Severity Index; ESS=Epworth Sleepiness Scale; PROMIS=Patient-Reported Outcomes Measurement Information System; SIDAS=Suicidal Ideation Attributes Scale; MEQ=Morningness-Eveningness Questionnaire; SOFAS=Social and Occupational Functioning Assessment Scale; ASRM=Altman Self-Rating Mania scale; WHODAS-2.0=WHO Disability Assessment Schedule 2.0; ReQoL-10=Recovering Quality of Life; WHO-ASSIST=WHO Alcohol, Smoking and Substance Involvement Screening Test; ASEC=Antidepressant Side Effect Checklist; DLMO=Dim light melatonin onset; EEG=Electroencephalography.

At the enrolment visit, participants will be given the opportunity to ask the Study Doctor and Clinical Research Manager (a nurse) any questions about the procedures and expectations of the trial. Subject to meeting eligibility criteria, participants will be asked to provide written informed consent (to the Study Doctor and/or Clinical Research Manager) and will then participate in a series of clinical, self-report, and medical history assessments. During the assessments, participants are offered comfort measures (e.g., tea, coffee), opportunities for breaks, and avenues for support in case of distress. They will be given an actigraphy watch (see **“**Biological and behavioural measures”) and sleep diary to wear and complete, respectively, for ∼2 weeks. During this period, participants who meet all the eligibility criteria will be randomised.

Participants will return ∼2 weeks later for the in-person baseline assessment (Table 1) and will receive the Investigational Product (melatonin/placebo or log-in for SHUTi). A subset of participants will undergo a physiological assessment of sleep and circadian rhythms before starting the intervention (see **“**Biological and behavioural measures”). During the 12-week intervention period, AEs and adherence will be monitored fortnightly and participants will complete follow-up assessments at Week 4 and 8 (via REDCap/MyCap for self-report and telephone for clinical assessments). To provide a safe and familiar participant experience, a member of the research team will be assigned to each participant to be a consistent contact and assessor throughout the trial. Subject to the Study Doctor’s discretion based on information gathered at the Week 4 assessment, participants in the medication arm will start taking an escalated dose (from 2mg to 4mg) starting at Week 5. Around the end of the 12-week intervention period, the trial team will schedule the end of treatment assessment visit (primary endpoint) and ∼6 months later, the team will schedule a post-discontinuation assessment.

### Primary and secondary outcomes

#### Primary efficacy endpoint

- Change in depressive symptoms from baseline to 12 weeks, as assessed by the QIDS-A-CR^62^

<u>Secondary endpoints</u> (*Note*: *common metric mandated by funder, **common metric recommended by consortium of teams with a 2023 Wellcome Mental Health Award):

- Partial remission of major depressive episode (MDE) at 12 weeks according to SCID-5^61^.
- Change in depressive symptoms from baseline to 12 weeks, assessed by the PHQ-9*^72^ and QIDS-A-SR^62^
- Change in anxiety symptoms from baseline to 12 weeks, assessed by the GAD-7*^73^ and the OASIS^74^
- Change in anxiety/depressive symptoms from baseline to 12 weeks, assessed by the RCADS*^75^ (18-year-olds only)
- Change in insomnia symptoms from baseline to 12 weeks, assessed by the ISI**^76^
- Change in general sleep-related impairment from baseline to 12 weeks, assessed by the PROMIS Sleep-Related Impairment short form 8a^63^
- Change in general sleep disturbance from baseline to 12 weeks, assessed by the PROMIS Sleep Disturbance short form^63^
- Change in chronotype from baseline to 12 weeks, assessed by the MEQ**^77^
- Change in quality of life from baseline to 12 weeks, assessed by the ReQoL^78^
- Change in social and occupational functioning from baseline to 12 weeks, assessed by the SOFAS^79^
- Change in disability from baseline to 12 weeks, assessed by the WHODAS 2.0*^80^
- Change in suicidal thoughts and behaviours from baseline to 12 weeks, assessed by the SIDAS^81^
- Change in hypo/manic symptoms from baseline to 12 weeks, as assessed by the ASRM^82^
- Change in alcohol, cannabis, and tobacco use from baseline to 12 weeks, assessed by the WHO-ASSIST^83^
- Change in sleep-waking timing from baseline to 12 weeks, assessed by the Consensus Sleep Diary*^84^
- Change in sleep-wake cycles from baseline to 12 weeks, assessed by actigraphy
- Change in dim-light melatonin onset (DLMO) time from baseline to 12 weeks, as measured by salivary melatonin sampling (20% subset of cases; see “Biological and behavioural measures” below)
- Change in sleep physiology (exploratory) from baseline to 12 weeks, assessed by Sleep Profiler^TM^ EEG Sleep Monitor (20% subset of cases; see “Biological and behavioural measures” below)

### Biological and behavioural measures

#### Genetics

All participants will be asked to provide a saliva sample for the purpose of investigating genetic predictors of treatment response (e.g., polygenic risk scores for sleep and circadian traits such as chronotype^86^). Samples will be analysed at the University of Queensland’s Human Studies Unit. The authors have experience collecting saliva for DNA from 500+ young people with mental disorders at the BMC^87–89^ and thousands of cases in other studies (e.g., Australian Genetics of Depression Study^90,91^, Australian Genetics of Bipolar Disorder Study^92^).

#### Actigraphy

All participants will be asked to wear a wrist-worn passive-sensing device (GENEActiv, Activinsights, UK) for 10-14 days (including two weekends) for the purpose of tracking whether the interventions cause changes in sleep-wake cycles and circadian rhythms. Actigraphy data will be collected before the intervention and near its completion (Table 1). Participants will be instructed to wear the device continuously on their non-dominant wrist, removing it only when showering, bathing, or swimming. The authors have experience collecting actigraphy data from 500+ young people with mental disorders at the BMC^87,93–95^ and ∼500 community-residing adolescents in Australia^96,97^.

#### Dim-light melatonin onset (DLMO)

A subset of participants (∼20%) will undergo melatonin sampling during chronobiology studies at the Woolcock Institute for Medical Research at Macquarie University for the purpose of testing whether the interventions cause changes to circadian rhythmicity of melatonin. This circadian assessment will be done before the intervention and near its completion (Table 1). Sampling will be performed according to established DLMO protocols^98^. Under dim-light (<5 lux) conditions and in a semi-recumbent position, hourly saliva samples will be collected from participants using passive drool kit, starting 6 hours before, and ending 2 hours after, their habitual bedtime (during which they are kept awake). Participants will be provided with non-electronic entertainment options while awake (e.g., music, audiobooks). Saliva samples will be analysed at the Adelaide Research Assay Facility. The authors have experience collecting overnight salivary melatonin samples in 70+ young people with mental disorders recruited at the BMC^99,100^.

#### Sleep physiology

The same subset (∼20%) of participants will undergo home-based assessments of sleep physiology. The Sleep Profiler^TM^ EEG Sleep Monitor (Advanced Brain Monitoring, Inc., USA) is a research-grade, wireless, multichannel forehead electroencephalography device that also records electrooculography, electromyography, pulse rate, head position, head movement, and snoring, and auto-stages several physiologic measures (e.g., sleep latency, sleep spindles, arousals). This assessment will be done before the intervention and near its completion (Table 1). The automated sleep staging of the Sleep Profiler^TM^ agrees strongly with human-scored polysomnography in clinical and nonclinical samples^101,102^. The device was chosen given its portability, ability to be self-applied, and therefore its feasibility in this large trial. The acceptability of the Sleep Profiler^TM^ in our population is not yet known.

#### Pupillometry

All participants will undergo an assessment to examine the pupil’s response to light (pupillary light reflex [PLR]), given the observation that the PLR can discriminate circadian from non-circadian forms of delayed sleep phase disorder^103^. The PLR will be measured by a PLR-3000 monocular pupillometer (NeurOptics, USA) in a windowless room (∼1-2 lux), using a protocol of 10-min of dark adaptation and two 1-sec light pulses (∼10 lux followed by ∼1500 lux). The authors have used this protocol in 100+ young people with mental disorders at the BMC^87^.

### Data management and security

Data collected for the purposes of the trial will be linked to unique study ID codes and will not contain identifiable information. Data collection will be conducted only by authorised members of the trial team, to whom this duty has been allocated, and who are named on the HREC application and Governance approvals for the trial. Research data will be stored in REDCap^68^ and on the University of Sydney’s hosted Research Data Store (RDS). The PI (IBH) and team (JJC, Clinical Research Manager [EJ]) will have access to the final trial dataset.

Publications or reports based on this trial will include only pooled results. Routine internal audits of data files will ensure completeness of data collection. Data for which hardcopies are generated will be stored securely in both original hardcopy and electronic form. Hardcopies will be retained so that comparison between electronic and original data is possible, to ensure accuracy of data entry and to resolve issues concerning any spurious data in the electronic file. These data will be kept under (1) lock and key at the trial site (BMC, University of Sydney) or (2) electronic file that is password-protected and accessible only by the trial team responsible for data entry or monitoring.

### Statistical Analysis Plan

The primary planned contrast on which the study is powered is the active intervention group (melatonin and dCBT-I arms) vs placebo. The secondary planned contrasts are melatonin vs dCBT-I; melatonin vs placebo; and dCBT-I vs placebo. A comprehensive data validation of the trial database will be performed before commencing analyses. A detailed final Statistical Analysis Plan (SAP) will be developed before database lock and unblinding, which will include prespecified estimands, sensitivity analyses, exact statistical models (e.g., covariance structure, adjustment for degrees of freedom), mock tables, etc, compliant within the R1 addendum to ICH E9^104^.

The primary outcome (QIDS-A-CR total) will be analysed using a linear mixed model for repeated measures (MMRM) including all available QIDS-A-CR scores (intention-to-treat) measured at weeks 4, 8, and 12 (end of treatment). Fixed effects will include the randomisation group, time point as a categorical variable, and the interaction between randomisation group and time point. The baseline QIDS-A-CR score will be included as a covariate alongside age and sex. The MMRM model will be used to estimate the effect of the intervention(s) at week 12 (end of treatment), expressed as the adjusted mean difference and its 95% confidence interval. Secondary outcomes will be analysed using a similar approach. For binary outcomes (e.g., MDE partial remission), logistic regression (binomial regression with logit link) will be used. Given that linear MMRMs use all data available and make valid inference under the assumption that data are Missing at Random, the primary analysis will not impute missing data. Broadly, sensitivity analyses will be conducted to assess the robustness of the results under different assumptions of the missing data mechanism (e.g., Missing Not at Random); these will be prespecified in the final SAP according to the R1 addendum to ICH E9^104^ prior to database lock and unblinding. Exploratory mediation analyses will be undertaken to examine whether changes in circadian rhythms (e.g., phase-advance of DLMO) mediate clinical changes (e.g., improvement in depressive symptoms). An interim analysis is not yet planned.

## CONCLUSION

The MELODY trial launched recruitment in August 2025. The trial aims to provide new knowledge about the value of melatonin and dCBT-I for improving the depressed phase of mood disorders in young people, and according to the interventionist model of causality^58^, the trial’s results may shed light on the nature of SCRDs as pathophysiological mechanisms of some types of mood disorders^31^.

## ETHICS AND DISSEMINATION

The MELODY trial will be performed according to the Declaration of Helsinki (2008) and the International Conference on Harmonisation–Good Clinical Practice (ICH-GCP) and has been reviewed and approved by the Human Research Ethics Committee (HREC) of the Sydney Local Health District (HREC Approval Number: X23-0450, Protocol version: 1.4, 7/7/25). The trial has been registered in the Australian New Zealand Clinical Trial Registry (ACTRN12624000017527). Any modifications will be submitted to the HREC for review prior to implementation as per HREC guidelines.

The findings of the trial will be disseminated into the scientific and clinical communities via peer-reviewed publications, talks, and clinician-facing outlets. LEWG will contribute to dissemination of the trial’s findings to the general public via relevant streams (e.g., social media, explainers, infographics). For each paper, all authors will satisfy the Vancouver criteria for authorship. There is no planned use of a professional writer.

## Trial sponsor

University of Sydney (within Australia: 1800 793 864; outside Australia: +61 2 8627 1444). The sponsor had no role in study design; collection, management, analysis, and interpretation of data; writing of the report; and the decision to submit the report for publication. The sponsor has no ultimate authority of any of these activities.

## Author contributions

JJC, SJH, NRW, JS, RRG, SLN, FI, KRM, SWC, SEM, YJCS, and IBH obtained funding for the trial. JJC wrote the first draft of the manuscript with input from MS and IBH. All authors contributed to subsequent drafts of the manuscript.

## Data statement

The datasets generated and/or analysed during this study will be accessible upon request to Prof Ian Hickie. The data provided will be fully anonymised, with all participant-identifiable information removed. Access to the full dataset will be granted only after the formal reporting of study findings in a peer-reviewed scientific journal. Datasets will be available exclusively to bona fide scientific researchers. Requests must be submitted in writing to the Principal Investigator, including details about the investigator’s background and the intended purpose of the data. Each request will be evaluated based on the proposed analyses, with potential uses likely to include meta-analyses, for example. The study’s Participant Information Sheet and consent forms explicitly mentioned the availability of anonymised data, a process approved by the Sydney Local Health District HREC.

## Acknowledgements

We are grateful to members of the Brain and Mind Centre Lived Experience Working Group for their help designing, planning, and conducting the trial. JJC is supported by a National Health and Medical Research Council (NHMRC) Emerging Leadership Fellowship (2008196). NRW is supported by a NHMRC L3 Leadership Fellowship (1173790). IBH is supported by a NHMRC L3 Leadership Fellowship (2016346).

